# Exploring the intersection between orthostatic hypotension and daytime sleepiness in Parkinson’s disease

**DOI:** 10.1101/2024.09.12.24313567

**Authors:** Abhimanyu Mahajan, Kevin R. Duque, Alok K. Dwivedi, Jesus Abanto, Luca Marsili, Emily J. Hill, Ameya Saraf, Kelsey J. McDonald, Adebukunola Arowosegbe, Heba A. Deraz, Aaron Bloemer, Alberto J. Espay

## Abstract

**Introduction:** Daytime sleepiness, reported in about 50% of patients with Parkinson’s disease (PD), is associated with high morbidity, poor quality of life and increased risk for accidents. While an association between dysautonomia and daytime sleepiness in early, de-novo PD has been reported, our understanding of the role of medications, cognitive status and co-morbidites on this relationship is inadequate.

**Methods:** Data were analyzed from the prospective Cincinnati Cohort Biomarkers Program. The primary outcome of interest was excessive daytime sleepiness (EDS), as measured by the Epworth Sleepiness Scale (ESS; ESS score > 10). The primary exposure variable was orthostatic hypotension (OH). Linear and logistic regression analyses followed by moderated graphical network analyses were conducted to explore the complex association between OH and ESS. Edge weight from graphical network analysis indicates the strength of the association.

**Results:** Data on 453 subjects with PD were analyzed. Median disease duration was 5.8 years and nearly 90% were H&Y stage <3. OH was not associated with EDS. OH was associated with depression (edge weight, 0.22) in cognitively impaired patients but not in cognitively normal patients. In addition, depression was associated with ESS (edge weight, 0.37; moderation weight, 0.22) in cognitively impaired patients to a greater extent than in cognitively normal patients (edge weight, 0.22).

**Conclusions:** OH is not directly associated with daytime sleepiness in early, treated PD. However, OH seems to be associated with ESS via depression in cognitively impaired patients. This complex relationship deserves additional study.

## 1. Introduction

Daytime sleepiness, difficulty in maintaining a desired level of wakefulness, is a disabling feature of neurodegenerative disorders^1^ and is noted in up to 50% of patients with Parkinson’s disease (PD).^2^ While it is associated with high morbidity, poor quality of life and increased risk for accidents in advanced PD, it may also be seen early in the disease raising the possibility of improvement in symptoms with appropriate intervention.^3^

Orthostatic hypotension (OH), secondary to cardiovascular dysautonomia, is present in approximately 50% of patients with PD. Even if asymptomatic, it may affect overall function and quality of life.^4^ On standing, when a drop in blood pressure is not accompanied by a corresponding increase in heart rate, it is referred to as neurogenic OH (nOH), a manifestation of sympathetic denervation in PD. OH has been associated with compromised mentation, with clinical observations suggesting a causal relationship through cerebral hypoperfusion.^5^ In healthy older adults, those with higher systolic blood pressure (BP) levels and diastolic BP variability during waking hours have been noted to have greater daytime sleepiness.^6^

Our understanding of this complicated relationship between OH and daytime sleepiness, especially the role of medications, comorbidities, and their impact on PD patients is inadequate. We sought to understand the link between these manifestations of PD with the goal of investigating if OH should be a potential therapeutic target to address daytime sleepiness in early, treated PD.

## 2. Material and methods

### 2.1 Study Sample

Data was prospectively collected as a part of the Cincinnati Cohort Biomarkers Program (CCBP).^7^ The CCBP systematically collects demographic and clinical data, including the use of validated scales, in patients with PD and other neurodegenerative disorders.^7^ Consecutive participants with a diagnosis of idiopathic PD^8^ and data on the primary exposure were included in this analysis. The study protocol was approved by the University of Cincinnati Institutional Review Board (IRB# 2018–5699).

### 2.2 Primary outcomes and exposures

Our primary outcome of interest was the Epworth Sleepiness Scale (ESS) score, a validated, self-reported measure of daytime sleepiness in PD.^9, 10^ Excessive daytime sleepiness (EDS) was defined with an ESS score > 10.^11^ The primary exposure variable was OH, defined by a decrease in systolic blood pressure (SBP) of ≥ 15 mmHg or diastolic blood pressure (DBP) of ≥ 7 mmHg, as previously validated from the sitting to the standing position^12^, which is the protocol for collecting orthostatic vital signs used in the CCBP. In the CCBP protocol, vitals are measured while sitting and after three minutes of standing. In addition, we performed sensitivity analysis by using a stricter cut-off of a decrease in SBP ≥ 20 mm Hg or DBP ≥10 mm Hg (revised OH or rOH), the standard lying-to-standing thresholds for OH.^13^ Another analysis using change in Mean Arterial Pressure (MAP, defined as diastolic BP + (1/3) * (systolic BP - diastolic BP) with posture was conducted to assess its effect on EDS and ESS scores. Finally, we also used as an exposure neurogenic OH (nOH), defined as a ratio of an increase in heart rate (HR) to a decrease in SBP of <0.5.^14^

### 2.3 Covariates

Demographic data: age, sex, education, patient-identified race (white or non-white) and ethnicity (non-Hispanic, Hispanic).

Clinical data: diagnosis, disease duration (time since motor symptom onset), Hoehn & Yahr (H&Y) stage, motor subscale of the Movement disorder society Unified Parkinson’s Disease Rating Scale (MDS UPDRS III), Montreal Cognitive Assessment (MoCA), Beck Depression Inventory (BDI), Beck Anxiety Inventory (BAI) and presence/absence of comorbidities, with particular interest in obstructive sleep apnea (OSA), Restless legs syndrome (RLS), anxiety, depression, cardiovascular disease, chronic kidney disease, cognition impairment, diabetes, and cancer. Anticholinergic burden (ACB), Gabapentin use and levodopa equivalent daily dose (LEDD) were extracted or computed from the data.^15, 16^

### 2.4 Data analysis

All scores and continuous variables were summarized with mean and standard deviation (SD) and non-normal data with median and interquartile range (IQR). Categorical variables were described with frequency and proportion. All baseline characteristics were compared with a t-test/Wilcoxon rank sum test or chi-square test according to OH and nOH status. In the unadjusted analysis, we compared ESS scores between exposure groups using unpaired t-tests while EDS between groups using a Chi-square test. Interactions between baseline characteristics and OH and nOH for the ESS outcome measure were also assessed using linear regression models. We performed adjusted analyses using a multiple linear regression model considering OH (or nOH) as the primary exposure variable. All prespecified covariates including age, sex, disease duration, education, number of comorbidities, ACB scores, LEDD, MDS-UPDRS III, H&Y stage, BDI, and BAI scores, were adjusted in the final analyses. Log-transformed LEDD, disease duration, MDS-UPDRS III, BDI, and BAI scores were used in the analyses. The adjusted analyses were further evaluated for EDS (ESS score > 10) using logistic regression models. Considering complex interactions between variables, we explored the association between OH and ESS in subgroups using linear and logistic regression models depending on the form of ESS outcome. We further evaluated the association of continuous markers of OH including MAP, drop in SBP, drop in DBP, and the ratio between increases in heart rate to decrease in SBP with EDS using logistic regression analyses. Considering the complex associations and moderating effects of cognitive impairment, we explored moderated network analysis or graphical network analysis to evaluate the intersections between OH and other variables for ESS.^17, 18^ The mixed graphical network analysis (MGM) employs the Ising-Gaussian model with an L1-regularized nodewise estimation method to estimate the pairwise strength of association. We used a cross-validation method to estimate pairwise estimates and moderation effects of cognition status as appropriate for small-sized studies. We used 500 bootstrap samples to determine the stability in the estimated edge weight and moderation weight and 95% confidence interval (CI). The MGM was employed separately for OH, rOH and nOH. We further performed a parsimonious MGM model by considering the most significant variables in the model. The final model was validated for stability in the estimated coefficients and 95%CI and reported with the MGM network graph. In the network graph, the gray line shows the paired association between a categorized variable with another variable while a green line indicates the paired positive association between continuous variables. The red lines indicate a negative paired association between continuous variables. The edge weight is interpreted as the standardized regression coefficient. The number indicated in the edge reports a proportion of nonzero effect across bootstrap samples. The results of linear regression analyses were described with regression coefficient (RC) while results of logistic regression with odds ratio (OR) with 95% CI and p-value. The results were considered statistically significant at a 5% level of significance. All statistical analyses were conducted using STATA 17 while MGM analysis was conducted with R4.4.1 using “mgm” package.

## 3. Results

A total of 453 PD subjects with data on the primary exposure and ESS variables were analyzed. Their mean age was 67.3 (SD: 9.5) years with a median disease duration of 5.8 (IQR: 3.3, 9.5) years. Nearly 90% of the subjects had H&Y stage <3. The mean MoCA score was 26.3 (SD: 3.2). Thirty-seven percent of patients had OH and 20.8% had nOH. Baseline characteristics between subjects with and without OH/ nOH were similar, except for higher age and disease duration in the OH cohort. (Table 1) There were no differences in the prevalence of OSA or RLS (p=0.36) between the two groups.

**Table 1.** Baseline characteristics of the cohort according to orthostatic hypotension and neurogenic orthostatic hypotension status.

| Factor | Entire cohort | non-OH | OH | p-value | non-nOH | nOH | p-value |
| --- | --- | --- | --- | --- | --- | --- | --- |
| **N** | 453 | 285 | 168 |  | 359 | 94 |  |
| Age (years), mean (SD) | 67.33 (9.47) | 66.20 (10.24) | 69.26 (7.64) | <0.001 | 66.68 (9.74) | 69.85 (7.87) | 0.004 |
| Sex-Male | 298 (65.9%) | 183 (64.4%) | 115 (68.5%) | 0.38 | 226 (63.1%) | 72 (76.6%) | 0.014 |
| Race-White | 437 (96.5%) | 272 (95.4%) | 165 (98.2%) | 0.12 | 346 (96.4%) | 91 (96.8%) | 0.84 |
| Ethnicity- Not Hispanic | 442 (99.1%) | 276 (98.6%) | 166 (100.0%) | 0.12 | 349 (98.9%) | 93 (100.0%) | 0.30 |
| Education-HS | 397 (89.8%) | 251 (90.3%) | 146 (89.0%) | 0.67 | 314 (89.7%) | 83 (90.2%) | 0.89 |
| Number of comorbidities |  |  |  | 0.62 |  |  | 0.93 |
| 0 | 151 (33.3%) | 100 (35.1%) | 51 (30.4%) |  | 120 (33.4%) | 31 (33.0%) |  |
| 1 | 184 (40.6%) | 111 (38.9%) | 73 (43.5%) |  | 147 (40.9%) | 37 (39.4%) |  |
| 2 | 64 (14.1%) | 43 (15.1%) | 21 (12.5%) |  | 51 (14.2%) | 13 (13.8%) |  |
| 3 | 32 (7.1%) | 20 (7.0%) | 12 (7.1%) |  | 25 (7.0%) | 7 (7.4%) |  |
| 4 | 15 (3.3%) | 7 (2.5%) | 8 (4.8%) |  | 10 (2.8%) | 5 (5.3%) |  |
| 5 | 6 (1.3%) | 3 (1.1%) | 3 (1.8%) |  | 5 (1.4%) | 1 (1.1%) |  |
| 6 | 1 (0.2%) | 1 (0.4%) | 0 (0.0%) |  | 1 (0.3%) | 0 (0.0%) |  |
| ACB score, mean (SD) | 0.60 (1.22) | 0.61 (1.22) | 0.59 (1.22) | 0.89 | 0.60 (1.19) | 0.61 (1.32) | 0.98 |
| LEDD, median (IQR) | 750.00 (400.00, 1125.00) | 685.00 (400.00, 1050.00) | 800.00 (450.00, 1200.00) | 0.15 | 750.00 (400.00, 1071.20) | 800.00 (575.00, 1200.00) | 0.14 |
| Disease duration, median (IQR) | 5.77 (3.26, 9.52) | 5.42 (2.92, 9.06) | 6.44 (3.67, 10.19) | 0.009 | 5.49 (3.14, 9.47) | 6.47 (4.34, 9.56) | 0.053 |
| MDS-UPDRS III, median (IQR) | 25.00 (17.00, 35.00) | 24.50 (17.00, 34.00) | 27.00 (18.00, 35.00) | 0.23 | 25.00 (17.00, 34.00) | 27.00 (19.00, 38.00) | 0.041 |
| H&Y Stage < 3 | 49 (11.0%) | 33 (11.7%) | 16 (9.6%) | 0.49 | 40 (11.3%) | 9 (9.7%) | 0.66 |
| BAI score, median (IQR) | 7.00 (4.00, 13.00) | 7.00 (3.00, 12.00) | 8.00 (4.00, 15.00) | 0.17 | 7.00 (4.00, 13.00) | 7.00 (3.00, 14.00) | 0.84 |
| BDI score, median (IQR) | 7.00 (4.00, 11.00) | 6.00 (3.00, 10.00) | 7.00 (4.00, 12.00) | 0.11 | 6.00 (3.00, 11.00) | 7.00 (4.00, 12.00) | 0.20 |
| MoCA score, mean (SD) | 26.27 (3.16) | 26.47 (3.19) | 25.93 (3.09) | 0.082 | 26.38 (3.13) | 25.87 (3.27) | 0.17 |
| MoCA<26 | 142 (31.5%) | 78 (27.4%) | 64 (38.6%) | 0.014 | 107 (29.8%) | 35 (38.0%) | 0.13 |

OH: orthostatic hypotension, SD: standard deviation; IQR: interquartile range, HS: high school, ABC: anti-cholinergic burden, LEDD: Levodopa Equivalent Daily Dose, MDS-UPDRS III: Movement Disorder Society Unified Parkinson's Disease Rating Scale, H&Y: Hoehn and Yahr, BAI: Beck Anxiety Inventory, BDI: Beck Depression Inventory, MoCA: Montreal Cognitive Assessment.

### 3.1 OH and EDS

There was no difference in mean ESS scores (7.2±5.0 vs. 6.7 ± 4.5, p=0.26) in PD subjects with and without OH (Supplementary data). The presence of OH was not associated with EDS (OR, 0.86, p=0.557) or ESS scores (RC −0.08; p=0.859) after adjusting for age, disease duration, sex, education, comorbidities, anticholinergic medication burden, LEDD, MDS-UPDRS III, H&Y stage, BAI, BDI, and MoCA score. Male sex (RC 1.1, p=0.015), BDI scores (RC 1.3, p<0.001), BAI scores (RC 0.8, p=0.008), and MoCA scores (RC −0.2, p=0.006) were significantly associated with EDS and ESS scores. (Supplementary data) There was no interaction between OH and other variables for ESS (Supplementary data).

Moderated network analysis showed that OH was associated with higher depression scores (edge weight, 0.10) in cognitively impaired patients (MoCA < 26)^19^ but not in cognitively normal patients. Moreover, the paired associations between depression and ESS (edge weight, 0.23; moderation weight=0.10) were more pronounced in cognitively impaired patients than in cognitively normal patients (edge weight, 0.10) (Supplementary data). The associations between OH and depression (edge weight, 0.16), and depression and ESS (edge weight, 0.37; moderation effect=0.22) did not change in the parsimonious adjusted models of cognitively impaired patients (Figure 1). The associations were found to be stable across multiple bootstrap samples and the moderating effect of cognitive impairment on the association between OH, depression, and ESS was significant. (Supplementary data)

**Figure 1.**
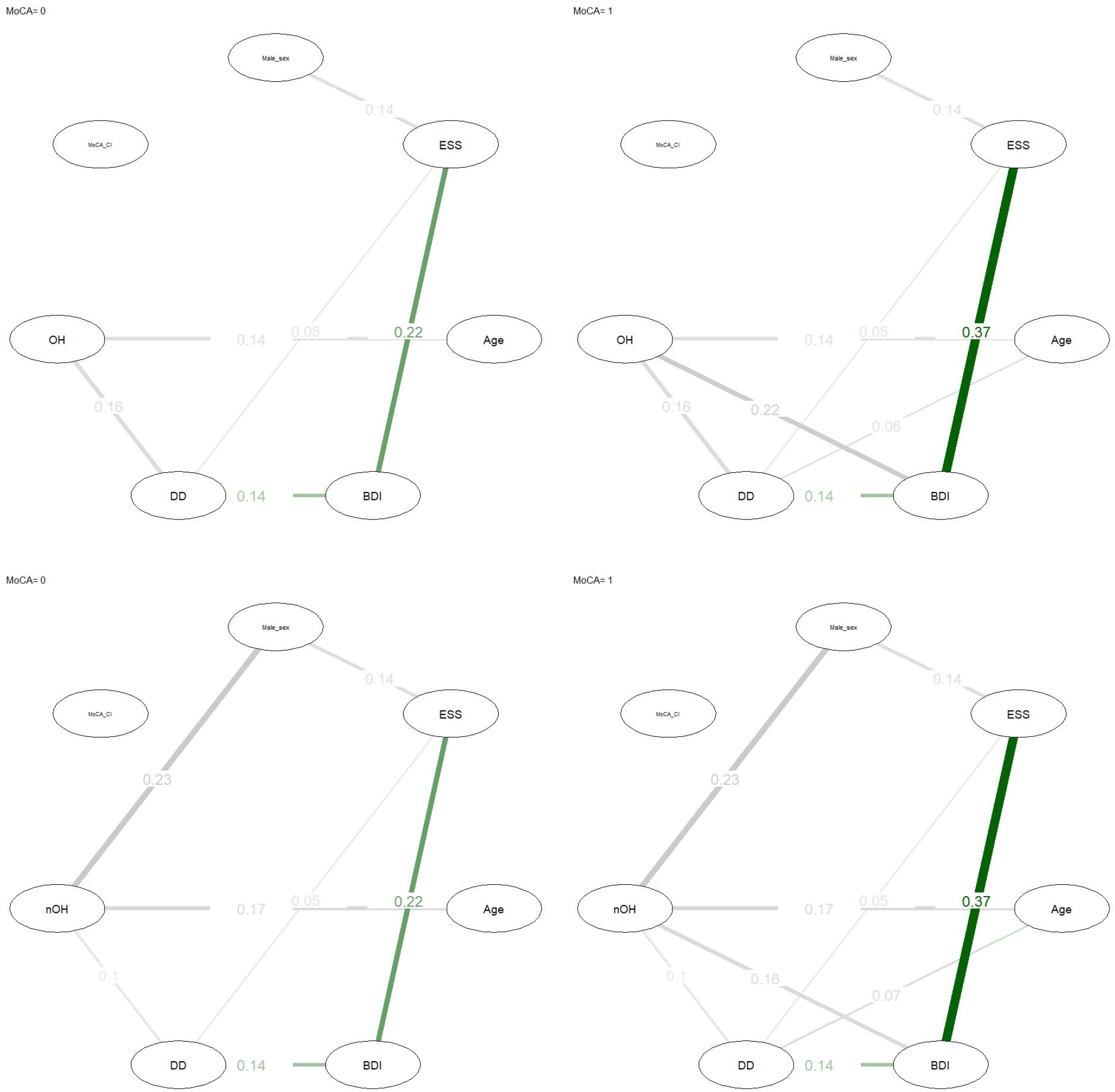
Moderated network analysis exploring the relationship between orthostatic hypotension and daytime sleepiness in cognitively impaired subjects with Parkinson’s disease

#### Sensitivity analyses

The stricter, rOH (cut off of a change in SBP ≤ 20 mm Hg or DBP ≤ 10 mm Hg from sitting to standing) was not associated with EDS (p=0.49) or ESS scores (p=0.46). (Supplementary data). Like OH, rOH was associated with depression (edge weight=0.26) and depression with ESS (edge weight=0.25, moderation weight=0.10) in cognitively impaired patients in network analysis (Supplementary data).

There was no statistically significant association between change in MAP with OH (p=0.71) or nOH (0.555). (Supplementary data)

### 3.2 nOH and EDS

Like OH, EDS (p=0.07) and ESS scores (p=0.1) were not significantly different according to nOH status in unadjusted analysis. (Supplementary data) nOH was not associated with EDS (OR, 1.24, =0.482) or ESS scores (RC, 0.18, p=0.734) in adjusted analyses. (Supplementary data) However, nOH was found to interact with multiple variables including education, UPDRS, stage, and depression. (Supplementary data)

In the moderated network analysis, nOH was only associated with depression (edge weight, 0.12) in cognitively impaired patients but not in cognitively normal individuals. In addition, depression was associated with ESS (edge weight, 0.22, moderation effect=0.10) in cognitively impaired patients to a greater extent than ESS (edge weight, 0.11) in cognitively normal patients. (Supplementary data) These associations were robust and highly statistically significant in models with reduced variables. (Figure 1 and Supplementary data)

## 4. Discussion

Our analysis of data from a clinically representative,^7, 20^ robust sample shows that OH is not associated with daytime sleepiness in PD. We confirmed these results by conducting analyses including BP as a continuous exposure variable (MAP) and ESS as a continuous outcome variables, thereby highlighting the absence of a relationship between BP and daytime sleepiness in early, treated PD.

The pathophysiology of daytime sleepiness is complex. Dopamine circuitry is central to the sleep-wake cycle and has been implicated in sleep issues seen in PD and restless leg syndrome.^2^ Sleep disturbances and daytime sleepiness are common non-motor features of PD, affecting nearly two-thirds of patients.^2, 21^ Dopamine dysfunction in the hypothalamus, a key structure in the regulation of the sleep-wake state, leading to direct/ indirect regulation of the orexinergic neurons has been hypothesized to be the underlying issue behind sleep issues in PD.^2^ OH has been shown to be associated with daytime sleepiness in advanced PD along with cognitive impairment and hallucinations.^22^ This would be consistent with the known contribution of underlying neurodegeneration to the autonomic system and sleep disruption,^2, 23^ with likely involvement of brainstem and reticular activating system in PD. The cholinergic system in PD is important to this discussion. A significant loss of cholinergic neurons in the brainstem have been reported in post-mortem studies of individuals with PD.^24^ These neurons innervate rostral and caudal targets including hypothalamic sleep-wake regulating regions. Severity of regional cholinergic neuron loss correlates with PD symptom severity.^25^ Our analysis is able to test the hypothesis of the contribution of OH to daytime sleepiness in early, treated PD without accompanying confounders and reports the absence of such a relationship.

A clinical correlation between excessive daytime sleepiness and dysautonomia in early, untreated PD has been previously reported.^11^ There were substantial similarities with study sample in terms of disease duration, motor severity (MDS UPDRS III), disease stage (H&Y), cognitive status, and sample composition by sex and race. Our approach differs as we account for medical comorbidities and medications (Anticholinergic medication use and LEDD), factors commonly encountered in clinic and important to our patient population. Our study sample was on a robust dose of dopaminergic medications (Median LEDD of 750 mg), suggesting that concerns for daytime sleepiness alone should not dissuade us from up titration of dopaminergic dose, if otherwise indicated. Our results additionally argue against targeting BP fluctuations as an approach to treat complaints of daytime sleepiness in early, treated, cognitively unimpaired PD.^26^

The clinical intersection of daytime sleepiness, OH and cognition in PD offers an interesting insight into underlying biological mechanisms. Daytime sleepiness may be seen in cognitively impaired PD patients with or without OH.^22^ OH is independently associated with cognitive impairment in PD. Early OH but not its symptom severity appears to increase the risk of dementia by 14% per year in PD.^27^ This association is not explained by more widespread Lewy, β-amyloid, tau, Alzheimer, or cerebrovascular pathologies. OH is known to be associated with significantly worse sustained attention, visuospatial and verbal memory in PD, and these findings are not explained by white matter disease burden.^28^ Locus coeruleus, implicated in both, noradrenaline release and attention, has also been proposed to explain the relationship between OH and cognitive impairment. Dopaminergic medications have a direct impact on OH in PD. It is possible that the reported impact of dopamine replacement on cognition may at least partially be secondary to the effect of OH on daytime sleepiness.^29^ Our analysis shows the absence of a direct relationship between OH and daytime sleepiness in an overall cognitively robust sample, thereby suggesting that daytime sleepiness may be construed as an expression of cognitive impairment in some PD patients. In those with cognitive impairment, our analysis indicated a more complex relationship between OH, depression and daytime sleepiness which requires a deeper study of the underlying pathophysiology, clinical manifestations and our measures. Underlying cognitive impairment may be a factor in determining this clinical presentation of OH.

OSA is common in PD and an important cause of daytime sleepiness. Our data showed no difference in the prevalence of OSA between those with and without OH. Similarly, no difference in the prevalence of RLS or Gabapentin use was noted between those with and without OH. Another strength of the study includes the use of comprehensive, prospectively collected data with a large sample size for analysis. Our patient population includes substantial representation of urban, suburban, and rural populations. Our analysis utilized established scales for assessment including the ESS, a well-known measure of daytime sleepiness. We conducted sensitivity analysis by utilizing a stricter cut-off for OH and a cut-off for nOH which did not alter our conclusions.

Our study is not without limitations. Our primary exposure is based on a cut-off that is not validated in PD. The assessment of vitals was done in the sitting and standing positions based on feedback of substantial discomfort with the tilt table by multiple subjects with PD in the CCBP program. Recognizing these issues, this approach has been deemed acceptable per recommendations of a consensus panel.^30^ We additionally performed analysis using modified, validated cut-offs to mitigate this shortcoming. Similarly, the ratio utilized for nOH has not been validated. The absence of a control group is another limitation of this analysis.

## 5. Conclusions

BP does not contribute to daytime sleepiness in early, treated PD. Daytime sleepiness alone should not be a deterrent against up titration of dopaminergic dose, if otherwise indicated. The nature of the relationship between BP and daytime sleepiness in the cognitive impaired needs additional, careful study.

## Data Availability

All data produced in the present study are available upon reasonable request to the authors

## 6. Acknowledgments

We want to thank all subjects who participated in the CCBP study.

## 7. Author’s contributions

AM: study conception and design, writing of the first draft of the manuscript, editing of final version of the manuscript

KRD: acquisition of data, revising the manuscript for content

AD: study design, data statistics and analysis, revising the manuscript for content JA: acquisition of data, revising the manuscript for content

LM: acquisition of data, revising the manuscript for content

EJH: acquisition of data, revising the manuscript for content

AS: acquisition of data, revising the manuscript for content

KJM: acquisition of data, revising the manuscript for content

AA: acquisition of data, revising the manuscript for content

HAD: acquisition of data, revising the manuscript for content

AB: acquisition of data, revising the manuscript for content

AJE: study conception, revising the manuscript for content, editing of final version of the manuscript

## 8. Financial disclosures

AM has received funding from the Dystonia Medical Research Foundation, Sunflower Parkinson’s disease Foundation and the Parkinson’s Foundation. He has served on the advisory board of Adaptive Biosciences through the Parkinson’s Study Group. He reports no conflicts of interest

KRD reports no financial disclosures

AKD reports no financial disclosures

JA reports no financial disclosures

LM has received honoraria from the International Association of Parkinsonism and Related Disorders (IAPRD) Society for social media and web support, and personal compensation as a consultant/scientific advisory board member for Acadia. Dr. Marsili has received a grant (collaborative research agreement) from the International Parkinson and Movement Disorders Society for the MDS-UTRS Validation Program (Role: PI), Non Profit.

EJH reports no financial disclosures

AS reports no financial disclosures

KM reports no financial disclosures

AA reports no financial disclosures

HAD has been funded by the Parkinson’s Foundation/ International Association of Parkinsonism and Related Disorders (IAPRD) outside of the submitted work.

AB reports no financial disclosures

AJE has received grant support from the NIH and the Michael J Fox Foundation; personal compensation as a consultant/scientific advisory board member for Mitsubishi Tanabe Pharma America (formerly, Neuroderm), Amneal, Acadia, Avion, Acorda, Kyowa Kirin, Supernus (formerly, USWorldMeds), NeuroDiagnostics, Inc (SYNAPS Dx), Intrance Medical Systems, Inc., Praxis Precision Medicines, Citrus Health, and Herantis Pharma; Data Safety Monitoring Board (chair) of AskBio; and publishing royalties from Lippincott Williams & Wilkins, Cambridge University Press, and Springer. He cofounded REGAIN Therapeutics and is co-inventor of the patent “Compositions and methods for treatment and/or prophylaxis of proteinopathies.”

## Notes

Declaration of interest: AM reports no relevant financial disclosures or conflicts of interest KRD reports no relevant financial disclosures or conflicts of interest AKD reports no relevant financial disclosures or conflicts of interest JA reports no relevant financial disclosures or conflicts of interest LM reports no relevant financial disclosures or conflicts of interest EJH reports no relevant financial disclosures or conflicts of interest AS reports no relevant financial disclosures or conflicts of interest KJM reports no relevant financial disclosures or conflicts of interest AA reports no relevant financial disclosures or conflicts of interest HAD reports no relevant financial disclosures or conflicts of interest AB reports no relevant financial disclosures or conflicts of interest AJE reports no relevant financial disclosures or conflicts of interest

### Competing Interest Statement

The authors have declared no competing interest.

### Funding Statement

This study did not receive any funding

### Author Declarations

The study protocol was approved by the University of Cincinnati Institutional Review Board

### Summary of Updates

Based on feedback, we have conducted additional analysis which led to additional insights and revised more nuanced conclusions. The older version needs to be updated to reflect this.

