## Supplementary data for "Exploring the intersection between orthostatic hypotension and daytime sleepiness in Parkinson’s disease"

**Supplementary Table 1.** Unadjusted differences in daytime sleepiness by orthostatic hypotension status

| **Factor** | **non-OH** | **OH** | **p-value** |
| --- | --- | --- | --- |
| N | 285 | 168 |  |
| ESS, mean (SD) | 6.71 (4.50) | 7.22 (4.95) | 0.26 |
| ESS>10 | 70 (24.6%) | 46 (27.4%) | 0.51 |
| **Factor** | **non-nOH** | **nOH** | **p-value** |
| N | 359 | 94 |  |
| ESS, mean (SD) | 6.71 (4.55) | 7.61 (5.10) | 0.1 |
| ESS>10 | 85 (23.7%) | 31 (33.0%) | 0.07 |
| **Factor** | **non-rOH** | **rOH** | **p-value** |
| N | 366 | 87 |  |
| ESS, mean (SD) | 6.83 (4.62) | 7.21 (4.91) | 0.49 |
| ESS>10 | 91 (24.9%) | 25 (28.7%) | 0.46 |

ESS: Epworth Sleepiness Score, SD; standard deviation, OH: orthostatic hypotension, nOH: neurogenic orthostatic hypotension, rOH: revised OH

**Supplementary Table 2.** Adjusted association of orthostatic hypotension and daytime sleepiness in Parkinson’s disease

|  | **Association between OH and ESS** | | | | **Association between nOH and ESS** | | | | |
| --- | --- | --- | --- | --- | --- | --- | --- | --- | --- |
| **Factors** | **RC** | **95%CI** | | **p-value** | **Factors** | **RC** | **95%CI** | | **p-value** |
| OH | -0.08 | -0.96 | 0.80 | 0.859 | nOH | 0.18 | -0.87 | 1.23 | 0.734 |
| Age | 0.02 | -0.03 | 0.06 | 0.513 | Age | 0.01 | -0.03 | 0.06 | 0.557 |
| Disease duration | 0.16 | -0.41 | 0.73 | 0.579 | Disease duration | 0.15 | -0.42 | 0.72 | 0.607 |
| Sex-male | 1.12 | 0.22 | 2.01 | **0.015** | Sex-male | 1.10 | 0.20 | 2.00 | **0.017** |
| Education-HS | -0.45 | -1.86 | 0.96 | 0.529 | Education-HS | -0.45 | -1.85 | 0.96 | 0.534 |
| Comorbidities | -0.01 | -0.38 | 0.37 | 0.974 | Comorbidities | -0.01 | -0.38 | 0.36 | 0.965 |
| ACB score | -0.08 | -0.43 | 0.27 | 0.658 | ACB score | -0.08 | -0.43 | 0.28 | 0.675 |
| LEDD | 0.13 | -0.08 | 0.33 | 0.217 | LEDD | 0.13 | -0.08 | 0.33 | 0.222 |
| MDS-UPDRS III score | 0.12 | -0.76 | 1.01 | 0.786 | MDS-UPDRS III score | 0.12 | -0.77 | 1.00 | 0.797 |
| H&Y Stage >2 | 0.29 | -1.11 | 1.70 | 0.681 | H&Y Stage≥2 | 0.31 | -1.09 | 1.72 | 0.66 |
| BAI score | 0.84 | 0.22 | 1.46 | **0.008** | BAI score | 0.85 | 0.23 | 1.47 | **0.007** |
| BDI score | 1.26 | 0.57 | 1.95 | **<0.001** | BDI score | 1.25 | 0.56 | 1.94 | **<0.001** |
| MoCA score | -0.20 | -0.35 | -0.06 | **0.006** | MoCA score | -0.20 | -0.35 | -0.06 | **0.006** |

ESS: Epworth Sleepiness Score, RC: regression coefficient, CI: confidence interval, OH: orthostatic hypotension, nOH: neurogenic orthostatic hypotension, DD: disease duration, ACB: anti-cholinergic burden, LEDD: Levodopa Equivalent Daily Dose, MDS-UPDRS III: Movement disorder society Unified Parkinson’s Disease Rating Scale, HnY: Hoehn and Yahr, BAI: Beck Anxiety Inventory, BDI: Beck Depression Inventory, MoCA: Montreal Cognitive Assessment. Log-transformed DD, LEDD, MDS-UPDRS III , BAI, and BDI scores were included in the analysis.

**Supplementary Table 3.** Adjusted associations of OH and nOH with abnormal daytime sleepiness (ESS>10)

|  | **Association between OH and ESS** | | | | **Association between nOH and ESS** | | | | |
| --- | --- | --- | --- | --- | --- | --- | --- | --- | --- |
| **Factors** | **OR** | **95%CI** | | **p-value** | **Factors** | **OR** | **95%CI** | | **p-value** |
| OH | 0.86 | 0.51 | 1.44 | 0.557 | nOH | 1.24 | 0.68 | 2.24 | 0.482 |
| Age | 1.01 | 0.98 | 1.04 | 0.382 | Age | 1.01 | 0.98 | 1.04 | 0.497 |
| Disease duration | 0.98 | 0.69 | 1.38 | 0.901 | Disease duration | 0.96 | 0.68 | 1.35 | 0.816 |
| Sex-male | 2.01 | 1.14 | 3.55 | **0.015** | Sex-male | 1.96 | 1.11 | 3.45 | **0.021** |
| Education-HS | 0.97 | 0.44 | 2.12 | 0.936 | Education-HS | 0.97 | 0.44 | 2.11 | 0.937 |
| Comorbidities | 1.08 | 0.88 | 1.33 | 0.463 | Comorbidities | 1.08 | 0.88 | 1.33 | 0.467 |
| ACB score | 0.98 | 0.80 | 1.19 | 0.819 | ACB score | 0.99 | 0.81 | 1.20 | 0.885 |
| LEDD | 1.08 | 0.95 | 1.23 | 0.247 | LEDD | 1.08 | 0.95 | 1.22 | 0.257 |
| MDS-UPDRS III score | 1.33 | 0.75 | 2.35 | 0.327 | MDS-UPDRS III score | 1.33 | 0.75 | 2.34 | 0.332 |
| H&Y Stage >2 | 0.51 | 0.22 | 1.18 | 0.117 | H&Y Stage≥2 | 0.53 | 0.23 | 1.21 | 0.13 |
| BAI score | 1.92 | 1.26 | 2.93 | **0.003** | BAI score | 1.93 | 1.26 | 2.95 | **0.002** |
| BDI score | 1.56 | 0.99 | 2.45 | **0.054** | BDI score | 1.53 | 0.97 | 2.40 | 0.067 |
| MoCA score | 0.90 | 0.83 | 0.98 | **0.014** | MoCA score | 0.91 | 0.84 | 0.98 | **0.015** |

ESS: Epworth Sleepiness Score, RC: regression coefficient, CI: confidence interval, OH: orthostatic hypotension, nOH: neurogenic orthostatic hypotension, DD: disease duration, ACB: anti-cholinergic burden, LEDD: Levodopa Equivalent Daily Dose, MDS-UPDRS III: Movement disorder society Unified Parkinson’s Disease Rating Scale, HnY: Hoehn and Yahr, BAI: Beck Anxiety Inventory, BDI: Beck Depression Inventory, MoCA: Montreal Cognitive Assessment. Log-transformed DD, LEDD, MDS-UPDRS III , BAI, and BDI scores were included in the analysis.

**Supplementary Table 4.** Factors interacting with the relationship between OH/ nOH and daytime sleepiness

|  | **With OH** | **With nOH** |
| --- | --- | --- |
|  | p-value | p-value |
| Age | 0.811 | 0.785 |
| Disease duration | 0.539 | 0.641 |
| Sex (Male) | 0.622 | 0.853 |
| Education (HS) | 0.330 | **0.043** |
| Comorbidities | 0.872 | 0.954 |
| ACB score | 0.659 | 0.715 |
| Anticholinergic medication use | 0.647 | 0.641 |
| LEDD | 0.952 | 0.682 |
| MoCA score | 0.985 | 0.467 |
| MDS-UPDRS III score | 0.590 | **0.056** |
| H&Y Stage (≥3) | 0.392 | **0.050** |
| BAI | 0.528 | 0.601 |
| BDI | 0.110 | **0.054** |

OH: orthostatic hypotension, nOH: neurogenic orthostatic hypotension, DD: disease duration, ACB: anti-cholinergic burden, LEDD: Levodopa Equivalent Daily Dose, MDS-UPDRS III: Movement disorder society Unified Parkinson’s Disease Rating Scale, H&Y: Hoehn and Yahr, BAI: Beck Anxiety Inventory, BDI: Beck Depression Inventory, MoCA: Montreal Cognitive Assessment.

**Supplementary Table 5.** Association of markers of orthostatic hypotension with daytime sleepiness in Parkinson’s disease

|  | OR | 95%CI | | p-value |
| --- | --- | --- | --- | --- |
| **Association between markers of OH and ESS>10** | | | | |
| MAP-sitting | 1.00 | 0.98 | 1.01 | 0.732 |
| MAP-standing | 0.99 | 0.97 | 1.01 | 0.283 |
| MAP-change | 1.01 | 0.99 | 1.04 | 0.271 |
| SBPdrop | 1.01 | 0.99 | 1.02 | 0.308 |
| DBPdrop | 1.01 | 0.99 | 1.04 | 0.339 |
| ∆HR/∆SBP | 0.98 | 0.92 | 1.04 | 0.555 |

ESS: Epworth Sleepiness Score, OR: odds ratio, CI: confidence interval, OH: orthostatic hypotension, nOH: neurogenic orthostatic hypotension, MAP: mean arterial pressure; SBP: systolic blood pressure; DBP: diastolic blood pressure; ∆: changes; DD: disease duration, ACB: anti-cholinergic burden, LEDD: Levodopa Equivalent Daily Dose, MDS-UPDRS III: Movement disorder society Unified Parkinson’s Disease Rating Scale, HnY: Hoehn and Yahr, BAI: Beck Anxiety Inventory, BDI: Beck Depression Inventory, MoCA: Montreal Cognitive Assessment.*Adjusted model included age, sex, education, number of comorbidities, disease duration, LEDD, ACB scores, UPDRS III, BAI, BDI, and MOCA. # Adjusted model included only significant variables (anxiety and sex). Log-transformed DD, LEDD, MDS-UPDRS III, BAI, and BDI scores were included in the analysis.
